# Evaluation of the Effect of Zinc, Quercetin, Bromelain and Vitamin C on COVID-19 Patients

**DOI:** 10.1101/2020.12.22.20245993

**Authors:** Amr Kamel, Heba Abdelseed, Yousef Albalawi, Eman Aslsalameen, Yousef Almutairi, Abdullah Alkattan

**Affiliations:** Tuberculosis Control Program, First Health Cluster, Ministry of Health, Riyadh, Saudi Arabia; Imam Abdulrahman Alfaisal Hospital, Riyadh, Saudi Arabia; Department of Pharmacy, King Khaled University Hospital, Medical City King Saud University, Riyadh, Saudi Arabia; Chronic Disease Prevention Center, Ministry of Health, Riyadh, Saudi Arabia; Department of Biomedical Sciences, College of Veterinary Medicine, King Faisal University, Al-Hofuf, 31982 Al-Ahsa, Saudi Arabia

**Keywords:** Quercetin, Bromelain, Zinc, Ascorbic acid, COVID-19, Inflammation

## Abstract

Coronavirus disease 2019 (COVID-19) is an infectious disease caused by a new strain of coronavirus. There are three phases of COVID-19: early infection stage, pulmonary stage and hyper-inflammation stage respectively. It is important to prevent lung or other organs injuries by preventing phase-II and phase-III via pharmacological or non-pharmacological treatments. This was a case series study done on twenty-two patients confirmed to be infected with SARS-CoV-2 and diagnosed with COVID-19. Patients in this study have been used quercetin 800 mg, bromelain 165 mg, zinc acetate 50 mg and ascorbic acid 1 g once daily as supplements for 3 to 5 days during SARS-CoV-2 infection. The aim of this study is to evaluate the safety and efficacy of quercetin, bromelain, zinc and ascorbic acid combination supplements on patients with COVID-19. The mean levels of WBC, ANC, ALC, AMC and AST were normal among all included patients before and after taking quercetin, bromelain, zinc and ascorbic acid supplements (P-value > 0.05). Quercetin 800 mg, bromelain 165 mg, zinc acetate 50 mg and ascorbic acid 1 g once daily supplements were safe for patients infected with SARS-CoV-2 and may prevent poor prognosis. Randomized clinical trials needed in the future to ensure the efficacy of quercetin, bromelain, zinc and vitamin c combination.

## Introduction

Coronavirus disease 2019 (COVID-19) is an infectious disease caused by a new strain of coronavirus.^[1]^ More than thirty million people around the world have been infected and got COVID-19, and at least one million of them died because of the disease complication including acute respiratory distress syndrome (ARDS) and cytokine storm.^[2]^

There are three phases of COVID-19: phase-I (early infection stage); in which SARS-CoV-2 starts to spread and proliferate and innate immunity activated. Phase-II (pulmonary stage); characterized by lung tissue injury and increased leucocytes recruitment. Phase-III (hyper-inflammation stage); which various organs could be damaged and there is an extreme exacerbation of immune response. In order to treat COVID-19 patients; it is important to prevent lung or other organs injury by preventing phase-II and phase-III via pharmacological or non-pharmacological treatments.^[3]^

Quercetin is a natural flavonoid molecule that distributed broadly in many fruits and vegetables including red onion, cranberry, kale, tomatoes, Hungarian wax and watercress.^[4]^ It was revealed in previous studies that quercetin has an anti-inflammatory and anti-hypersensitivity effect by preventing pro-inflammatory prostaglandins and leukotrienes through inhibiting of cyclooxygenase (COX) and lipoxygenase (LOX) enzymes, therefore; quercetin was used as an extract in various trials to treat different infectious and non-infectious diseases.^[5]^ In addition, quercetin showed to reduce tumor necrosis factor-alpha (TNF-α) production with chronic inflammation.^[6]^ Reduction in the ratio of CD4^+^:CD8^+^ T cells and suppression of macrophages, dendritic, mast cells and interleukin-6 (IL-6) levels were revealed after a specific tissue was treated with quercetin in pre-clinical studies (see figure.1).^[7-8]^ Besides, quercetin expected to has antiviral activity by acting as a zinc chelator and as a zinc ionophore as well.^[9]^ However, because most of these studies were done by using quercetin in-vitro with high concentration and in-vivo studies cannot use the same doses; it’s showed minimum effect during clinical trials. The available data clarifies that quercetin is a very safe molecule and used as a nutritional supplement with a dose reached 1500 mg divided per day.^[10]^

**Figure.1.**
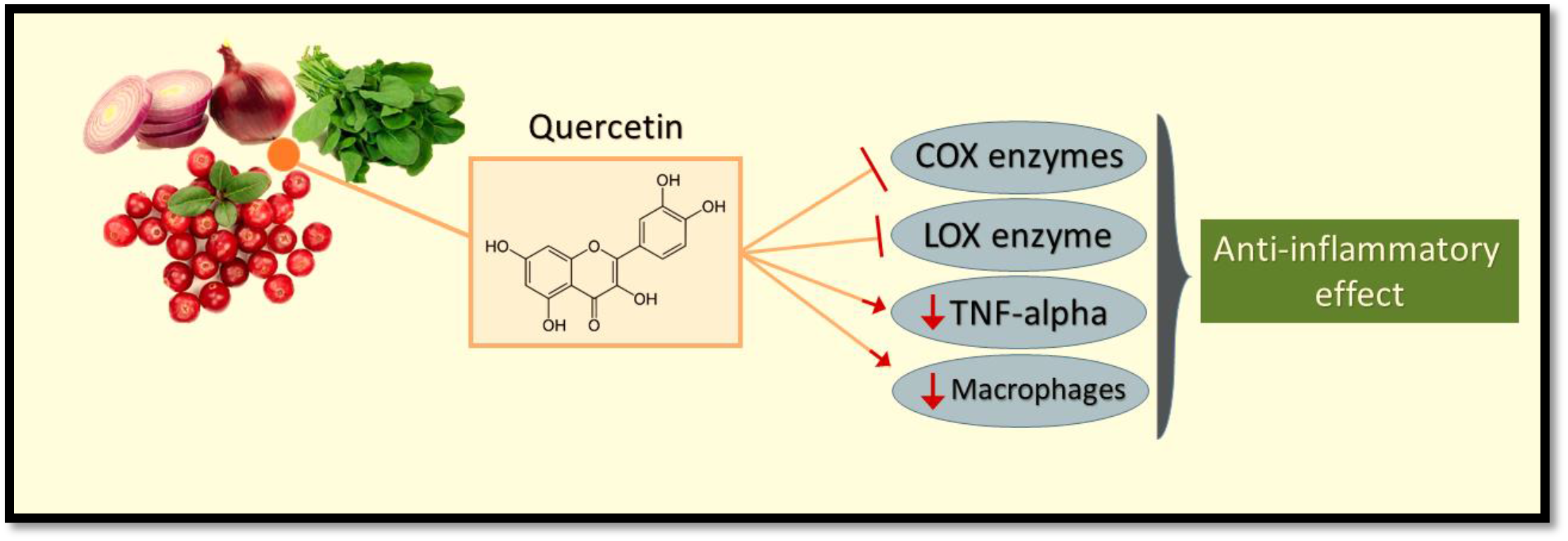
Role of quercetin in inhibiting inflammation by blocking the activity of COX (cyclooxygenase) enzymes and LOX (lipoxygenase) enzyme, in addition to reducing TNF-α (tumor necrosis factor-alpha) and macrophages levels.

Bromelain is a protein enzyme that found mainly in the stem of the pineapple plant. The bioavailability of bromelain was high through the oral route, and was safe even when consumed more than 11 grams per day.^[11]^ In vitro studies showed that bromelain exerts anti-inflammatory effects through reducing bradykinin serum^[12]^ and modulating the expression of some genes related to inflammation.^[13]^ Three genes related to inflammation including TLR4, TNF-α and IL-8 were found to be less expressed after bromelain treatment (see figure.2). On the other hand, PPARγ gene expression was elevated after treatment with bromelain.^[14]^ Therefore, bromelain may have a role in reducing inflammations during various disorders and may be used in combination with other analgesics and anti-inflammatory drugs.

**Figure.2.**
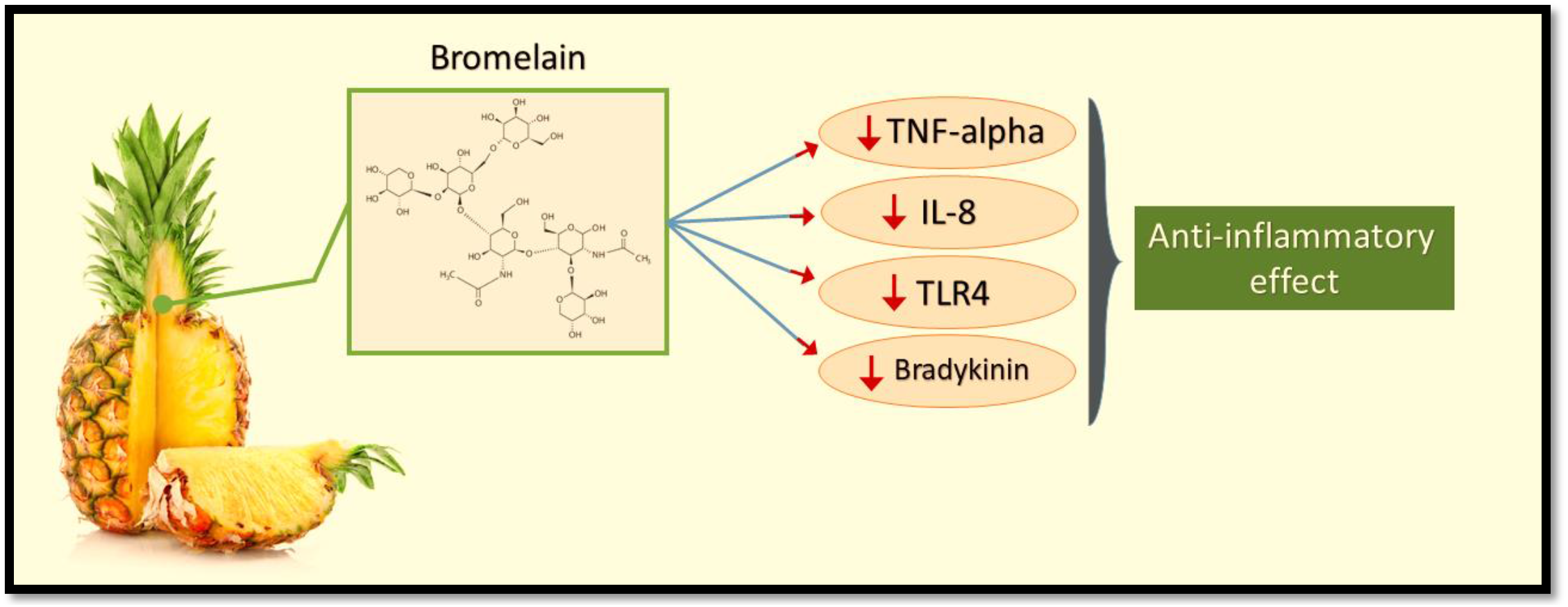
Role of bromelain in preventing inflammation by lowering of TNF-α (tumor necrosis factor-alpha), IL-8 (interleukin-8), TLR4 (toll-like receptor-4) and bradykinin levels.

Since the inflammatory status of patients during COVID-19 may lead to severe consequences and even death if not prevented or treated adequately; it is important to ensure high quality care to patients and provide evidence-based prophylaxis and treatment.

The aim of this study is to evaluate the efficacy of quercetin, bromelain, zinc and ascorbic acid supplements on patients with COVID-19.

## Methods

### Study design and subjects

This was a case series study conducted between June and September 2020 on twenty-two patients confirmed to be infected with SARS-CoV-2 and diagnosed with COVID-19. The study subjects included are adults and hospitalized in Imam Abdulrahman Alfaisal Hospital in Riyadh.

### Supplements and measurements

COVID-19 patients in this study have been used quercetin 800 mg, bromelain 165 mg, zinc acetate 50 mg and ascorbic acid 1 g once daily as supplements for 3 to 5 days during SARS-CoV-2 infection. A number of laboratory tests were done for all patients included in this study. These tests include absolute neutrophil count (ANC), absolute lymphocyte count (ALC), absolute monocyte count (AMC), hemoglobin (Hb), platelets (Plts), potassium (K), aspartate aminotransferase (AST), oxygen saturation percentage (SaO2), D-dimer and white blood cells (WBCs). In addition, medical and medication history were reported.

### Endpoint and statistical analysis

The primary endpoint was to ensure the efficacy of quercetin, bromelain, zinc and ascorbic acid supplements by evaluating the laboratory results pre- and post-supplements. Paired t-test was used to reveal the differences between different lab tests before and after quercetin, bromelain, zinc and vitamin c combination supplement among patients.

### Ethical approval

Institutional review board (IRB) was obtained from the Saudi ministry of health on the 7^th^ of June 2020 with the central IRB log number: 20-95M.

## Results

Twenty-two hospitalized patients diagnosed with COVID-19 were enrolled in this study, all of them were taking quercetin, bromelain, zinc and ascorbic acid as supplements. The mean age of patients was 49.27 years, and 59% of them were older than 50 years old. The percentage of male patients was 68.18%. More than 13% of the patients were having chronic diseases. About half of the patients were receiving antibacterial and antiviral medications during hospitalization, and 63.63% of total patients were on anti-coagulants. Days of stay average was 9 days (see table.1).

**Table.1.** Patients’ Baseline Characteristics.

| Variables | All COVID-19 cases<br>(N=22) |
| --- | --- |
| Mean age (in years) | 49.27 |
| Older than 50 years (%) | 59.09 |
| Male (%) | 68.18 |
| Diabetes (%) | 22.72 |
| Hypertension (%) | 13.63 |
| Antibacterial use (%) | 54.54 |
| Antiviral use (%) | 40.9 |
| Anti-platelet or anti-coagulant use (%) | 63.63 |
| Mean Days of hospital stay | 9 |

The mean D-dimer level at admission was elevated (1.0082 mcg/ml). Mean WBCs levels at admission and at discharge were 7440 and 8550 cells/mm^3^ respectively (P-value = 0.34). Mean ANC at admission and at discharge were 5570 and 5800 cells/microliter respectively (P-value = 0.86). O2sat% mean was less than 94% at admission, and was more than 94% at discharge (P-value = 0.83). AST mean levels were slightly elevated at admission and at discharge (46 and 44.8 U/L respectively, P-value = 0.9). Mean ALC was 1240 at admission and was 1740 cells/microliter at discharge (P-value = 0.11). Mean platelets count at admission and at discharge were 243830 and 304200 cells/microliter respectively (P-value = 0.45). The mean AMC was 456 at admission and 587 cells/microliters (P-value = 0.09). Regarding hemoglobin mean levels, it was 13.68 at admission and 13.24 g/dl at discharge (P-value = 0.78). Mean potassium concentration at admission and at discharge were 4.53 and 4.38 mmol/l (P-value = 0.45) (see table.2).

**Table.2.** Laboratory Tests Pre- and Post-Supplements.

| Lab tests<br>-mean values- | Pre-supplements<br>(N=22) | Post-supplements<br>(N=22) | P-value |
| --- | --- | --- | --- |
| WBCs (cells/mm3) | 7440 | 8550 | 0.34 |
| Hb (g/dl) | 13.68 | 13.24 | 0.78 |
| K (mmol/L) | 4.53 | 4.38 | 0.45 |
| Platelets<br>(cells/microliter) | 243.83 | 304.2 | 0.45 |
| AST (Units/L) | 46.08 | 44.8 | 0.9 |
| ANC<br>(cells/microliter) | 5.57 | 5.8 | 0.86 |
| ALC<br>(cells/microliter) | 1.24 | 1.74 | 0.11 |
| AMC<br>(cells/microliter) | 0.46 | 0.59 | 0.09 |

## Discussion

Quercetin supplement was and still interested by many researchers globally since various types of studies were focusing on it. Regarding studies about infectious diseases; quercetin was studied with Zika virus^[15]^, Ebola virus^[16]^, murine coronavirus^[17]^, dengue virus^[18]^, SARS-CoV-2^[19]^ and influenza A virus^[20]^, and most of these studies conclude that quercetin may have a substantial role as prophylactic or treatment of different types of viruses. Unlike quercetin, bromelain supplement was not widely studied about its efficacy against infections, however, few types of researches done claimed that bromelain could prevent or eradicate some microorganisms including Escherichia coli^[21]^ and SARS-CoV-2.^[22]^

In this study, the mean D-dimer level of patients diagnosed with COVID-19 was more than 0.5 mcg/ml; which indicates that their condition was not mild and need hospitalization based on the Chinese study. In addition to quercetin and bromelain supplements, most of the twenty-two patients were on hospital medications which include vitamin C, zinc, enoxaparin, drugs expected to have an anti-SARS-CoV-2 effect (ribavirin, hydroxychloroquine or lopinavir-ritonavir) and antibacterial drugs. As shown in the results, all the patients’ lab tests done at admission and at discharge were not significantly different and the mean days of stay at the hospital was 9 days. These results reveal that quercetin 800 mg once daily with bromelain 165 mg, in addition to zinc acetate 50 mg and vitamin c 1 g supplements are safe with COVID-19 patients who were on multiple therapies including antivirals and antibacterial medications. The efficacy of quercetin, bromelain, zinc and ascorbic acid combination was not clear in this study, because of lacking placebo or comparable group; however, their efficacy in preventing severe consequences of SARS-CoV-2 infections cannot be ruled out based on previous studies (see figure.3). Large comparable studies need to be done about quercetin and bromelain to confirm their efficacy in treating COVID-19 cases.

**Figure.3.**
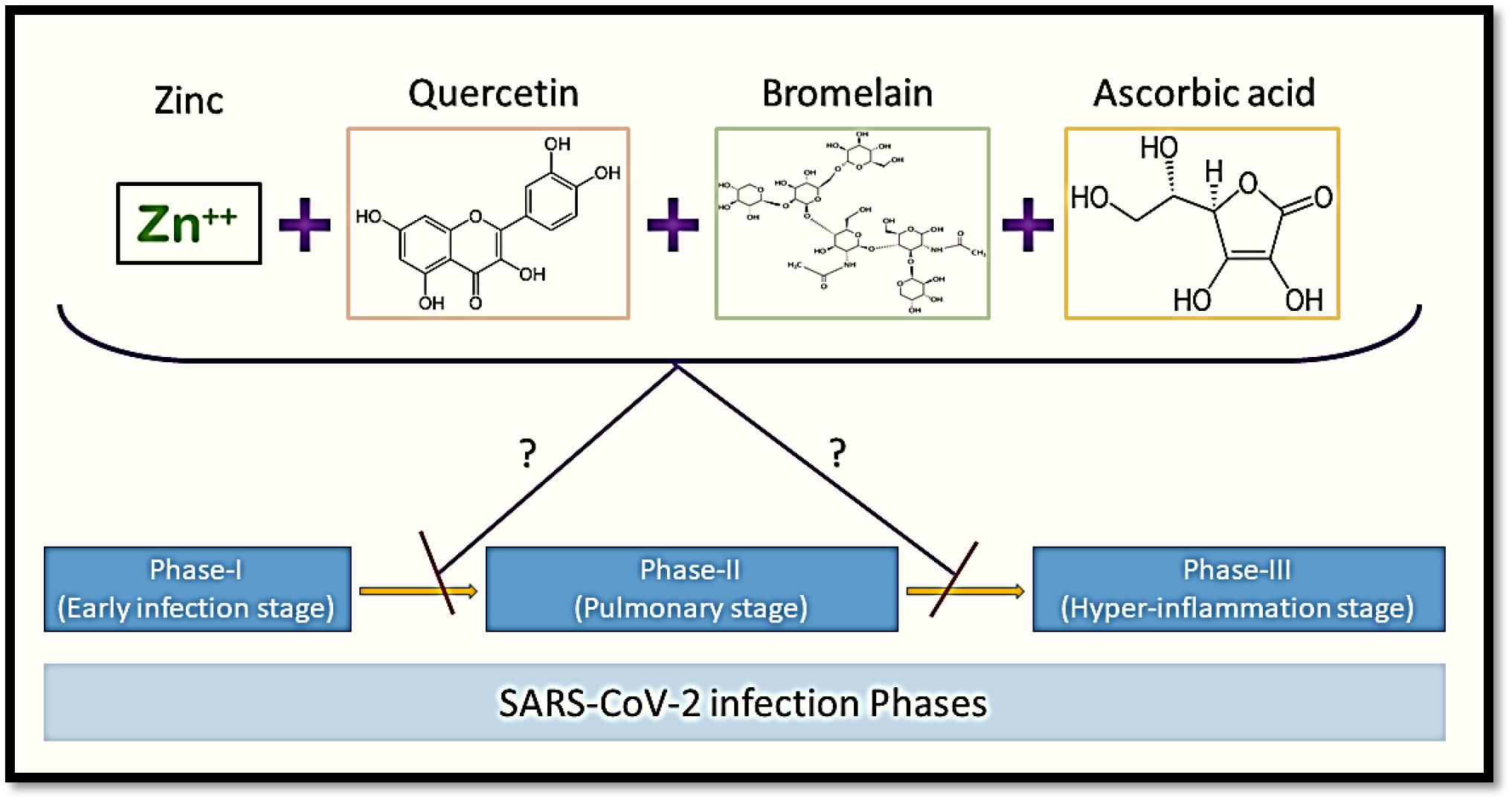
The expected efficacy of quercetin, bromelain, zinc and ascorbic acid combination in preventing poor prognosis of COVID-19 patients by restraining from pulmonary and hyper-inflammation stages.

## Conclusion

Quercetin 800 mg, bromelain 165 mg, zinc acetate 50 mg and ascorbic acid 1 g once daily as supplements for 3 to 5 days were safe for patients infected with SARS-CoV-2 and may prevent poor prognosis through restraining from hyper-inflammation and cytokine storm. Randomized clinical trials are needed in the future to ensure the efficacy of quercetin, bromelain, zinc and ascorbic acid combination.

## Data Availability

All data confidentially saved and available in special hard ware and used only by the research's members.

## Conflict of interest

The authors have no conflict of interest.

## Notes

### Competing Interest Statement

The authors have declared no competing interest.

### Clinical Trial

NCT04468139

### Author Declarations

Approval letter was given from the central institutional review board for research ethics of the Ministry of Health, Saudi Arabia, with IRB log no. 20-95M for one year starting from 7th of June, 2020. Decision made: Approved letter was given (Approved).

